# Combined oral contraceptive use and serotonin 2A and 2C receptor brain architecture in healthy women

**DOI:** 10.1101/2025.07.08.25331039

**Authors:** Anna Kauffmann, Anjali Sankar, Vincent Beliveau, Claus Svarer, Brice Ozenne, Patrick MacDonald Fisher, Vibe G. Frokjaer, Søren Vinther Larsen

## Abstract

**Objectives:** Combined oral contraceptive (COC) use is linked to increased depression risk, potentially via serotonergic pathways. This study examined whether serotonin 2A/2C receptor (5-HT2AR/5-HT2CR) brain binding differs between healthy women using COCs and non-users.

**Methods:** 71 healthy women had been scanned with either [18F]Altanserin (17 COC users, 22 non-users) or [11C]Cimbi-36 Positron Emission Tomography (17 COC users, 15 non-users). Multiple linear regression and latent variable models were used to assess associations between COC use and neocortical 5-HT2AR and subcortical-HT2AR/5-HT2CR binding, respectively.. Analyses were performed on data pooled across both radiotracers and on each tracer, separately.

**Results:** In pooled analyses across both tracers, COC use was not significantly associated with 5-HT2AR binding in the neocortex (−7.7%, 95% CI [−18.9;5.2], p=0.22), nor with 5-HT2AR/5-HT2CR in subcortical regions (−7.8, 95% CI [−21.7;7.7], p=0.31). In [11C]Cimbi-36-only analyses, COC use was associated with −12.6% (95% CI [−22.1;−1.9], p=0.02) lower 5-HT2AR binding in neocortex and −23.5% lower 5-HT2AR/5-HT2CR binding in subcortical regions (95% CI [−35.6;-9.1], p=0.002). No significant differences were observed in the [18F]Altanserin-only analyses.

**Conclusion:** The [11C]Cimbi-36 data indicated lower cortical 5-HT2AR and subcortical 5-HT2AR/5-HT2CR binding in COC users compared to non-users, but this was not observed in the [18F]Altanserin data. This may reflect better signal-to-noise properties of [11C]Cimbi-36 and the fact that it binds more selectively to the high-affinity, biologically active receptor state. These results offer potential mechanistic insights into the depression risk associated with COC use and may have implications for treatments targeting 5-HT2AR/5-HT2CR, underscoring the need for replication and further investigation.

**Highlights:**

1) COC use was associated with lower 5-HT2AR/CR brain binding in Cimbi-36 PET data
2) COC use was not associated with 5-HT2AR brain binding in [18F]Altanserin PET data
3) We speculate if lower 5-HT2AR/CR levels may affect treatments targeting 5-HT2AR/CR

## Introduction

Combined oral contraceptives (COCs) are used by approximately 151 million women globally and represent one of the most commonly used forms of hormonal contraception in Denmark (United Nations, 2019), (Lidegaard et al., 2009). COCs consist of two steroid hormones; typically, a synthetic estrogen paired with a synthetic progestogen. Also, COCs with natural estrogen exist, however they are very rarely used. These contraceptives primarily work by suppressing ovulation through inhibition of the hypothalamic–pituitary–gonadal axis, leading to reduced levels of endogenous ovarian sex steroids. In addition to their contraceptive function, COCs are frequently prescribed for the treatment of gynecological disorders such as dysmenorrhea and endometriosis. However, their use has been associated with potential adverse effects, including an increased risk of thromboembolic events (Lidegaard et al., 2009), breast cancer (Mørch et al., 2017) and mental health concerns as several large-scale, population-based studies have reported an association between the initiation of hormonal contraceptives - including COCs - and a heightened risk of developing depression (Zettermark et al., 2018),(Skovlund et al., 2016),(Johansson et al., 2023),(Larsen et al., 2025). Despite these consistent findings, the biological mechanisms that may explain an increased risk for depression remain poorly understood.

Sex steroid receptors are widely distributed throughout the brain (Barth et al., 2015), which allows synthetic hormones in COCs to influence brain function both directly and indirectly, through the suppression of natural ovarian hormone levels; one study suggests that the dose of synthetic progestogen in hormone-releasing intrauterine devices correlates with the risk of depression, pointing to a potential direct effect of the synthetic progestogen (levonorgestrel) (Larsen et al., 2024); prior work from our group using a pharmacological model of sex hormone suppression demonstrated a plausible causal link between suppressed natural ovarian hormone levels and the emergence of depressive symptoms, likely mediated through alterations in serotonin signaling in the brain (Frokjaer et al., 2015). Both estrogen and progestogen are known to influence serotonergic pathways (Barth et al., 2015); estrogen, in particular, has been shown to modulate brain serotonin metabolism (Bethea et al., 2002),(Barth et al., 2016),(Barth et al., 2015), increase the availability of serotonin transporters (Lu et al., 2003),(Sumner et al., 2007),(Suda et al., 2008), and thereby affect overall serotonergic neurotransmission. Similarly, progestogen can influence the expression of genes and proteins involved in serotonin signaling (Barth et al., 2015).

Interestingly prior work of ours also points to potential effects of COC use on serotonin receptor levels: COC use was associated with a lower serotonin 4 receptor (5-HT4R) brain level in healthy women (Larsen et al., 2020), a pattern that mirrors findings in patients diagnosed with depression (Köhler-Forsberg et al., 2023). Thus, these results support the hypothesis that alterations in the brain’s serotonin system may contribute to the increased risk of depression observed in COC users.

Other components of the serotonergic system, including the serotonin 2A receptor (5-HT2AR), are also sensitive to sex steroids. This receptor is abundantly expressed in the brain (Barth et al., 2015), particularly in the neocortex (Beliveau et al., 2017). Three studies have shown that treatment with estrogen replacement therapy in postmenopausal women significantly increased the 5-HT2AR binding by about 37% in the prefrontal cortex and that the 5-HT2AR level was positively associated with plasma estradiol levels (Kugaya et al., 2003),(Moses-Kolko et al., 2003), (Moses-Kolko et al., 2011). A small cross-sectional positron emission tomography (PET) study from our group investigated 5-HT2AR binding in healthy women using the 5-HT2AR antagonist radiotracer, [18F]Altanserin, and found no evidence of a difference (−0.02 (05% CI [−0.27–0.22], p=0.84) in cortical 5-HT2AR binding between healthy women using COCs vs. non-users (Frokjaer et al., 2009). However, the study included only 29 participants (14 COC users) and may have been limited to rule out a potential small to moderate effect. In the present study, we include these original participants along with additional 10 individuals (total n = 39) scanned with [18F]Altanserin PET available from the Center for Integrated Molecular Brain Imaging (Cimbi) database (Knudsen et al., 2016).

More recently, the Cimbi database also has expanded to include a dataset of 5-HT2AR PET scans with the [11C]Cimbi-36 radiotracer - a 5-HT2AR and serotonin 2C receptor (5-HT2CR) agonist with high test–retest reliability (Ettrup et al., 2016). Of the two receptors, the 5-HT2AR is the dominant receptor in neocortex, whereas they are both expressed subcortically (Finnema et al., 2014). Compared to [18F]Altanserin, [11C]Cimbi-36 provides improved signal-to-noise ratio, binds to biologically active receptors, and may offer better signal in subcortical regions. This enables a more nuanced investigation of the potential effects of COC use on 5-HT2AR availability in the neocortex and 5-HT2AR/5-HT2CR availability in subcortical regions.

Therefore, the present study aimed to assess the effects of COC use on 5-HT2AR/5-HT2CR binding in healthy women. Our primary objective was to compare neocortical 5-HT2AR levels between COC users and non-users. As secondary objective, we evaluated 5-HT2AR/5-HT2CR binding in subcortical regions including the hippocampus, putamen, caudate nucleus, thalamus, and amygdala.

## Materials and methods

### Participants

Data were available from the Cimbi database (Knudsen et al., 2016). They comprised data on 98 PET brain scans from healthy women conducted with one of two radiotracers: [18F]Altanserin or [11C]Cimbi-36 who were recruited for brain imaging studies between 2001-2020 at the Neurobiological Research Unit at Rigshospitalet in Copenhagen, Denmark. The Ethical Committee in the capital region of Denmark approved all study protocols ((KF)02-058/99), (KF)01-156/04, (KF)01-001/02, (KF)11-061/03, (KF)01-156/04, KF-01-124/04, (KF)01-2006-20, (KF)01-2006-20, H-4-2012-105, H-16026898)).

Women included in the studies had no current or previous psychiatric or primary somatic disorders and had standard physical and neurological examinations. Of these, women were excluded based on the following criteria: 1) age 50 or older, or self-reported postmenopausal status [n=19]; 2) use of estrogen-containing treatments other than COCs (e.g., Estrogel and Estrofem) [n=2]; 3) use of other forms of hormonal contraception, such as intrauterine hormonal devices and progestogen-only pills [n = 2]; 4) unknown hormonal contraceptive status [n=2]; 5) withdrawal from COC use within two months prior to the PET scan [n=1]; 6) missing data on relevant covariates [n=1]. In total, 71 healthy women were included in the analyses, of which 32 were scanned using the [11C]Cimbi-36 radiotracer and 39 using the [18F]Altanserin radiotracer. Of the 39 women scanned with [18F]Altanserin, 29 were also included in the study by Frokjaer et al. (2009). A detailed flowchart outlining participant selection from the Cimbi database is provided in the Supplementary Material (**Figure S1**).

Participants were screened for depressive symptoms with the Major Depression Inventory (MDI) questionnaire (Bech et al., 2001). Educational attainment was measured using a 5-point Likert scale, where a score of 1 indicated no vocational qualification and a score of 5 represented completion of more than four years of higher academic education. Personality traits, specifically neuroticism, were assessed using the NEO Personality Inventory–Revised (NEO PI-R) (Hansen et al, 2005). The neuroticism score consists of six facets: 1) anxiety, 2) hostility, 3) depression, 4) self-consciousness, 5) impulsiveness, and 6) vulnerability to stress. A composite subscore was derived from the inward-directed facets - anxiety, depression, self-consciousness, and vulnerability to stress - as previous research has demonstrated a positive correlation between this subscore and frontolimbic 5-HT2AR binding (Frokjaer et al., 2008),(Høgsted et al., 2025).

### Combined oral contraceptive user status

Women were asked about their hormonal contraceptive user status through face-to-face interviews on the day of the PET scan or written questionnaire specifically about COC use. Women with missing information from the face-to-face interview, but who did not indicate use of hormonal contraception when asked about current medication use or who answered “no” specifically to COC use on the written questionnaire were classified as non-users (n=13) in the main analyses. However, for sensitivity analyses, these women were considered to have uncertain hormonal contraceptive status and analysed separately.

### Brain imaging

High-resolution structural T1-weighted magnetic resonance images were acquired on either a Siemens 1.5 Tesla Vision scanner (Erlangen, DE) (n=13), a Siemens 3-Tesla Magnetom Trio scanner (n=26), a Siemens 3 T Magnetom Verio scanner (n=15), or a Siemens 3 T Prisma scanner (n=17). The T1-weighted image was used for segmentation into gray matter, white matter, and cerebrospinal fluid, and for delineation of regions of interest (ROIs). [18F]Altanserin- and [11C]Cimbi-36 PET images were co-registered with the T1-weighted image to obtain the binding potentials of the ROIs. The primary ROI was the neocortex due to the high expression of 5-HT2AR in the neocortex and the high signal-to-noise ratio (Beliveau et al., 2017). Secondary ROIs included subcortical regions; the hippocampus, amygdala, caudate, putamen, and thalamus, as we had not previously studied these regions in this regard (Frokjaer et al., 2009).

The PET scans were obtained with an 18-ring GE-Advance scanner (General Electric, Milwaukee, WI, USA) (n=39) or a Siemens ECAT High-Resolution Research Tomograph (HRRT) (CTI/Siemens, Knoxville, TN, USA) (n=32). All [18F]Altanserin scans were acquired using the GE-Advance scanner, and all [11C]Cimbi-36 scans were acquired using the HRRT scanner. Both scanners operated in three-dimensional acquisition mode with an in-plane resolution of approximately 6 mm and <2 mm (Ettrup et al., 2016) respectively. The [18F]Altanserin PET scans were performed according to the method described by Pinborg et al. (Pinborg et al., 2003). This protocol includes an [18F]Altanserin bolus injection followed by a continuous infusion to attain tracer steady-state conditions in the brain and blood. The scans consisted of a 10-minute transmission scan followed by 40 minutes (4×10min) of dynamic PET-scan acquisition, starting two hours after the bolus injection when steady state conditions were achieved. Venous blood samples were drawn during the scan and corrected for radiolabeled metabolites of [18F]Altanserin to measure the concentration of free [18F]Altanserin in plasma. The acquired data were reconstructed using a filtered back projection algorithm followed by attenuation-, dead time-, and scatter correction. The [11C]Cimbi-36-PET scans were performed according to the method described by Ettrup et al. (Ettrup et al., 2014). The scans included a 6-minute transmission scan followed by 120 minutes (frames: 6 × 10 s, 6 × 20 s, 6 × 60 s, 8 × 2 min, and 19 × 5 min) dynamic PET scan. Data were reconstructed using a 3D-OSEM-PSF algorithm with a point spread function (PSF) of 4 mm (Comtat et al., 2008).

In line with established methods (Knudsen et al., 2016), motion correction was conducted using the AIR algorithm developed by Woods, Cherry, and Mazziotta (Woods et al., 1992). PET images underwent within-frame Gaussian smoothing, with a kernel size of 12 mm for the GE-Advance scanner and 10 mm for the HRRT scanner. For image co-registration for the [18F]Altanserin scans, a semi-automatic visual overlay method was used, based on in-house software, Mars (Willendrup et al., 2004), whereas for [11C]Cimbi-36 scans, the co-registration was performed via a fully automated method using SPM8. The [18F]Altanserin plasma corrected binding potential (BP_P_) was calculated as a brain-to-blood ratio at steady state and the [11C]Cimbi-36 non-displaceable binding potential (BP_ND_) was quantified by using a simplified reference tissue model using the cerebellum as a reference region (Ettrup et al., 2014),(Pinborg et al., 2003). If the coefficient of variance for a regional binding potential was larger than ±100%, or if a regional binding potential was negative, the kinetic modeling was considered unsuccessful, why such regional binding potentials were considered missing. This was the case for putamen in 7 cases, for caudate in 3 cases, and for amygdala in 2 cases. 5-HT2AR BP_P_ for [18F]Altanserin and 5-HT2AR BP_ND_ for [11C]Cimbi-36 are considered indices of the 5-HT2AR density, but specifically subcortical [11C]Cimbi-36 BP_ND_ is considered indices of 5-HT2AR/5-HT2CR density (Finnema et al., 2014). [11C]Cimbi-36 and [18F]Altanserin show a high correlation across cortical regions (mean Pearson’s r: 0.95 ± 0.04); thus, pooling data across the two tracers is considered reasonable (Ettrup et al., 2016). However, due to potential differences in subcortical binding, analyses of subcortical areas should be interpreted with caution.

For illustrative purposes, we generated vertex-wise cortical 5-HT2AR BP_ND_ maps to visualize the distribution of cortical 5-HT2AR BP_ND_ differences between the COC users and non-users. This illustration was limited to the neocortex, where a single average value from a large pooled region was used in the regression analyses.

The vertex-wise cortical 5-HT2AR BP_ND_ maps were generated using the PetSurfer (Greve et al., 2014) processing stream in FreeSurfer version 7.4.1 (http://surfer.nmr.mgh.harvard.edu) (Fischl, 2012). Briefly, PET-MR co-registration was estimated using linear registration between the structural MRI and the time-weighted average of the dynamic PET images. Cortical vertex-wise time activity curves (TACs) were obtained by resampling the dynamic PET data onto the common average surface space (fsaverage) registration (Postelnicu et al., 2009),(Fischl et al., 1999). Cortical TACs were surface smoothed by 10 mm full width at half-maximum. Kinetic modelling of the parametric BP_ND_ was performed using the Multilinear Reference Tissue Model 2 (MRTM2) (Ichise et al., 2003) with cerebellar grey matter (GM), as reference region. Cerebellar GM masks were obtained by taking the intersection of the cerebellar GM segmentation defined by PetSurfer and a cerebellar GM segmentation, excluding vermis, derived using SUIT 3.1 (Diedrichsen, 2006). The reference region washout rate (k2’) was computed using MRTM (Ichise et al., 1996), and the high-binding TAC was obtained from a surface-weighted average of the neocortical regions. Percent difference maps were generated by fitting our multiple linear regression model (described below) at each vertex using log-transformed BP_ND_. The regression coefficients were back-transformed to percent difference using the formula **(e^β^ − 1)* 100**.

### Statistics

We used multiple linear regression models to model the association between log-transformed neocortical 5-HT2AR binding and COC use adjusted for age, body mass index (BMI), neuroticism subscore, radiotracer, and magnetic resonance (MR) field strength (1.5T vs. 3T). Assuming homoscedasticity (i.e., constant variance of the errors across all levels of the independent variables), the back-transformed COC coefficient can be interpreted as the percent differences in mean 5-HT2AR levels between COC users and non-users. Given that most of the [¹⁸F]Altanserin data were previously reported (Frokjaer et al., 2009), that [¹¹C]Cimbi-36 offers improved signal-to-noise ratio in subcortical regions (Ettrup et al., 2016), and that the association of COC use with 5-HT2AR binding has not previously been examined using [¹¹C]Cimbi-36, we conducted a [¹¹C]Cimbi-36-only analysis to explore whether similar associations could be observed using this tracer. This analysis involved a multiple linear regression model adjusted for age, BMI, and neuroticism subscore. Furthermore, injected [11C]Cimbi-36 per kg bodyweight was considered as a potential confounder, as it was observed to differ between the groups. However, since it was administered in tracer doses and therefore accounted for in the kinetic modeling, and since it was not associated with BPND, it was not included in the model.

For our secondary objective, we assessed the association between COC use and subcortical 5-HT2AR/5-HT2CR binding by using a linear latent variable model (LVM). Here, the effect of COC use was evaluated as mediated through the shared covariance across the hippocampus, amygdala, caudate, putamen, and thalamus. The log-transformed binding potentials were adjusted for age, BMI, neuroticism subscore, radiotracer, and MR field strength when using data from both radiotracers. When restricting the analysis to [¹¹C]Cimbi-36, adjustments included age, BMI, and neuroticism subscore only. Percent differences in regional 5-HT2AR/5-HT2CR levels between COC users and non-users are reported after performing back-transformation. To handle missing data of subcortical binding potentials, the LVMs were estimated using full information maximum likelihood (FIML) which leads to consistent estimates when subcortical binding potentials are missing at random (Holst and Budtz-Jørgensen, 2013). Furthermore, a sensitivity analysis was conducted in which we excluded women with uncertain hormonal contraceptive status. Lastly, for comparison we also provide [¹⁸F]Altanserin-only analyses largely resembling the previous analyses (Frokjaer et al., 2009).

Diagnostic tools were used to assess the adequacy of the multiple linear regression models’ assumptions. Score tests were used to detect possible misspecification in the covariance parameters in the LVM: the covariance parameter with the smallest p-value was added to the LVM if any test showed a Benjamini-Hochberg adjusted p-value below 0.05. This was performed recursively and using only individuals with complete data (n=61 for the pooled analysis and n=23 for the Cimbi-36-only analysis) instead of under FIML due to software limitation. As visual inspection of residuals appeared to deviate from a normal distribution in the analyses on neocortex and on the pooled analysis on subcortical regions, we also provide sensitivity analyses using median regression for these. For the LVM, a latent factor model was used to summarize the binding into a single latent outcome which was then regressed against COC use in the median regression. We used two-sided statistical tests where p-values less than 0.05 were considered statistically significant. Statistical analyses were performed in R version 4.3.0 (R Core Team, 2023), and the lava package was used to create the LVM (Holst and Budtz-Jørgensen, 2013). The study was pre-registered at https://aspredicted.org/ (113493) before data was obtained from the Cimbi database.

## Results

Of the 71 participants, 34 were COC users, and 37 were non-users. The study population’s demographics, psychometrics, and PET parameters are presented in **Table 1**. The COC users were younger than the non-users (median age (1^st^ quantile, 3^rd^ quantile) of 25.4 (27.0, 39.1) years vs. 33.0 (21.6, 28.3) years), but otherwise, the groups were well balanced across BMI, neuroticism subscore, education score, and MDI. Median (1^st^ quantile, 3^rd^ quantile) MDI scores of 5.0 (4.0, 5.0) and 4.0 (3.0, 5.0) indicates generally low levels of depressive symptoms across the groups, although one scored as high as 27 and 26 in each group, but were not diagnosed with depression following a clinical interview evaluation.

**Table 1.** Clinical profiles and PET parameters.

| **Table 1. Clinical profiles and PET parameters** |  |  |  |
| --- | --- | --- | --- |
|  | **Non-users (n=37)** | **COC users (n=34)** | **n** |
| **Clinical profiles** |  |  |  |
|  | **Median (Q1, Q3)** | **Median (Q1, Q3)** |  |
|  | **[Range]** | **[min, max]** |  |
| Age | 33.0 (27.0, 39.1)<br>[19.1, 49.7] | 25.4 (21.6, 28.3)<br>[19.0, 44.7] | 71 |
| BMI [kg/m<sup>2</sup>] | 24.2 (21.7, 26.6)<br>[19.3, 45.9] | 23.0 (21.3, 26.2)<br>[18.4, 43.1] | 71 |
| Neuroticism subscore<sup>a</sup> | 45.0 (38.0, 58.0)<br>[10.0, 94.0] | 53.0 (40.0, 59.8)<br>[19.0, 104.0] | 71 |
| Education score | 4.5 (3.0, 5.0)<br>[1.0, 5.0] | 5.0 (4.0, 5.0)<br>[1.0, 5.0] | 64 |
| MDI | 4.0 (3.0, 7.0)<br>[0.0, 26.0] | 5.0 (2.0, 7.0)<br>[0.0, 27.0] | 66 |
| **PET parameters** |  |  |  |
|  | **n (%)** | **n (%)** |  |
| Individuals scanned with [11C]Cimbi-36-PET | 15 (40.5) | 17 (50.0) | 71 |
| Individuals with 3T MR scan | 30 (81.1) | 28 (82.4) | 71 |
|  | **Median (Q1, Q3)** | **Median (Q1, Q3)** |  |
|  | **[min, max]** | **[min, max]** |  |
| Injected dose [MBq] ([18F]Altanserin) | 241.0 (196.0, 309.5)<br>[143.0, 437.0] | 266.0 (213.5, 302.0)<br>[155.0, 447.0] | 34 |
| Injected dose [MBq] ([11C]Cimbi-36) | 576.8 (391.1, 596.7)<br>[213.0, 604.0] | 498.0 (454.9, 569.0)<br>[238.0, 600.0] | 32 |
| Inj. mass per kg [µg] ([18F]Altanserin) | 0.023 (0.017, 0.034)<br>[0.004, 0.127] | 0.033 (0.025, 0.050)<br>[0.010, 0.134] | 24 |
| Inj. mass per kg [µg] ([11C]Cimbi-36) | 0.006 (0.003, 0.011)<br>[0.001, 0.026] | 0.013 (0.008, 0.019)<br>[0.004, 0.024] | 32 |
| Specific radioactivity [GBq/µmol]<br>([18F]Altanserin) | 82.9 (59.6, 145.9)<br>[12.0, 358.0] | 48.1 (36.0, 75.5)<br>[11.3, 155.9] | 27 |
| Specific radioactivity [GBq/µmol] ([11C]Cimbi-36) | 583.6 (322.4, 750.0)<br>[73.4, 1274.0] | 189.4 (131.1, 497.3)<br>[95.4, 645.0] | 32 |
| Plasma [18F]Altanserin [kBq/ml] | 2.1 (2.1, 2.2)<br>[2.0, 2.7] | 2.1 (2.1, 2.2)<br>[2.0, 2.2] | 37 |
| Free fraction [18F]Altanserin [%] | 0.3 (0.3, 0.4)<br>[0.2, 1.4] | 0.4 (0.3, 0.5)<br>[0.2, 0.9] | 39 |
| Cerebellum AUC [kBq/ml] ([11C]Cimbi-36) | 2163.6 (1953.6, 2591.0)<br>[1803.5, 4909.0] | 2071.6 (1954.7, 2408.1)<br>[1688.4, 3301.8] | 22 |

<sup>a</sup>\*Neuroticism subscore: consisting of four out of six facets of the neuroticism score from the NEO Personality
Inventory-Revised.
COC: combined oral contraceptive, SD: standard deviation, BMI: body mass index, MDI: Major Depressive Inventory,
AUC: Area Under the Curve (standardized to injected dose per kilogram of body weight).

The median specific radioactivity at the time of injection was lower and the median injected [11C]Cimbi-36 mass per kg bodyweight were higher in the COC users group compared to in the non-users group. Accordingly, they had received comparable injected radioactive dosages and the areas under the curves (AUC) in the cerebellar reference region in the analyses with the [11C]Cimbi-36 radiotracer were comparable between groups, indicating similar free and non-displaceable signals between groups. Last, the groups had a similar distribution of MR scanner types with different field strengths (82.2% of the non-users vs. 82.4% of the COC users were scanned with a 3T MR scanner).

### 5-HT2AR binding in neocortex

In the analysis based on the pooled data across the [18F]Altanserin and the [11C]Cimbi-36 PET radiotracers, we found no evidence of a difference in 5-HT2AR binding between COC users and non-users in neocortex (−7.7%, 95% CI [−18.9;5.2], p=0.22) (**Figure 1A**). However, when restricting our analysis to [11C]Cimbi-36-only scans, we found −12.6% (95% CI [−22.1;−1.9], p=0.02) lower BP_ND_ in COC users compared to non-users (**Figure 1B**). This effect in neocortex is illustrated in **Figure 2** based on vertex-based parametric images. Sensitivity analyses, - excluding women with uncertain hormonal contraceptive status and median regression analyses, are presented in **Table S1**. The result of these were largely consistent with the main findings, except COC users showed even lower binding levels when excluding women with uncertain hormonal contraceptive user status. In line with the previous study(Frokjaer et al., 2009), we found no evidence of a difference between the groups when only using the [18F]Altanserin scans (−5.0%, 95% CI [−24.5;19.6], p=0.66) (**Figure S2**).

**Figure 1.**
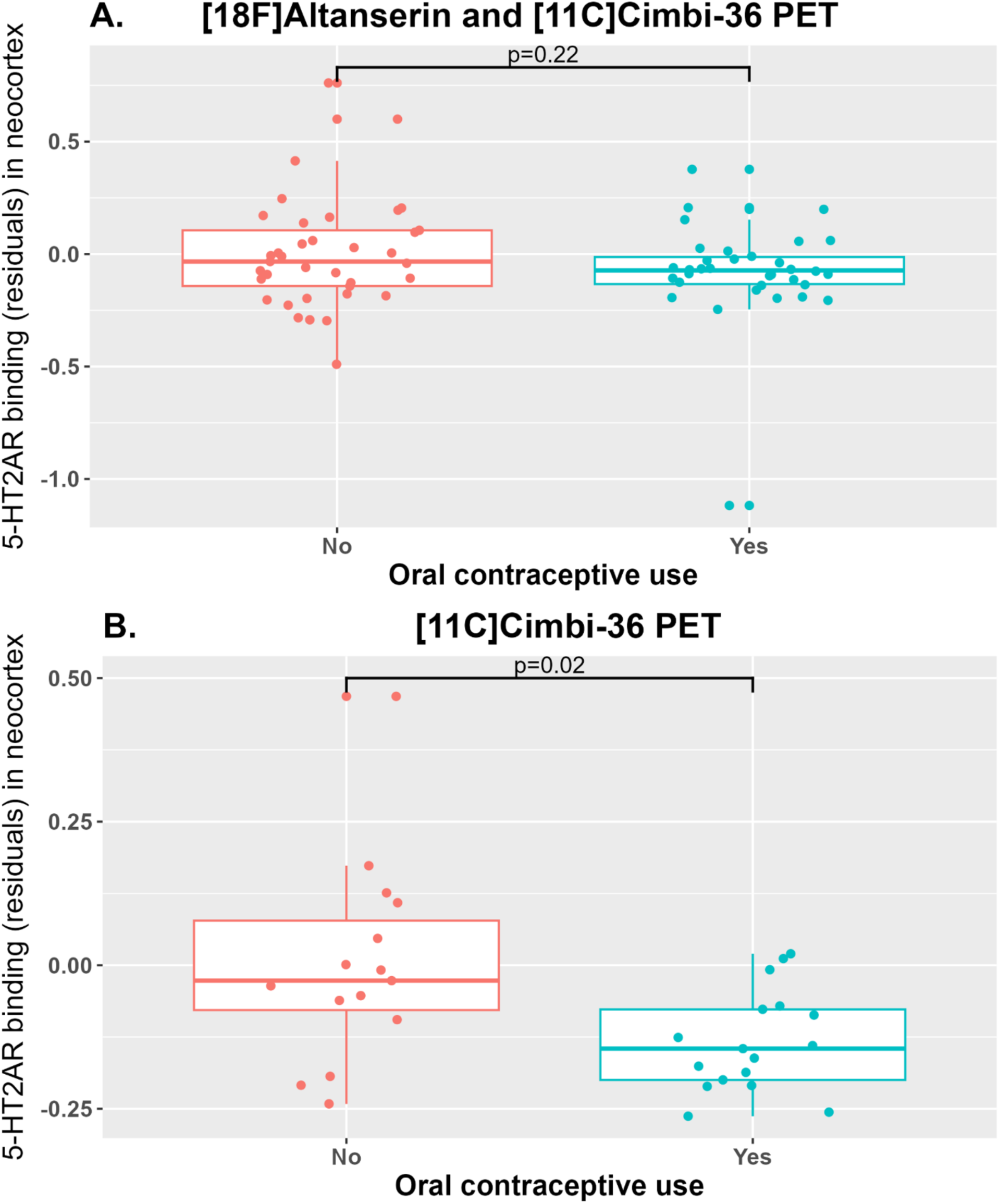
Partial residual plot of log-transformed 5-HT2AR binding potentials in neocortex in COC users and non-users (**A**) [18F]Altanserin and [11C]Cimbi-36 radiotracer scans and (**B**) in [11C]Cimbi-36-only scans. P-values were computed with multiple linear regression models adjusted for mean-centered age, body mass index, neuroticism subscore, and for the pooled analysis also radiotracer variant ([11C]Cimbi-36 as reference) and magnetic resonance field strength (3T as reference).

**Figure 2.**
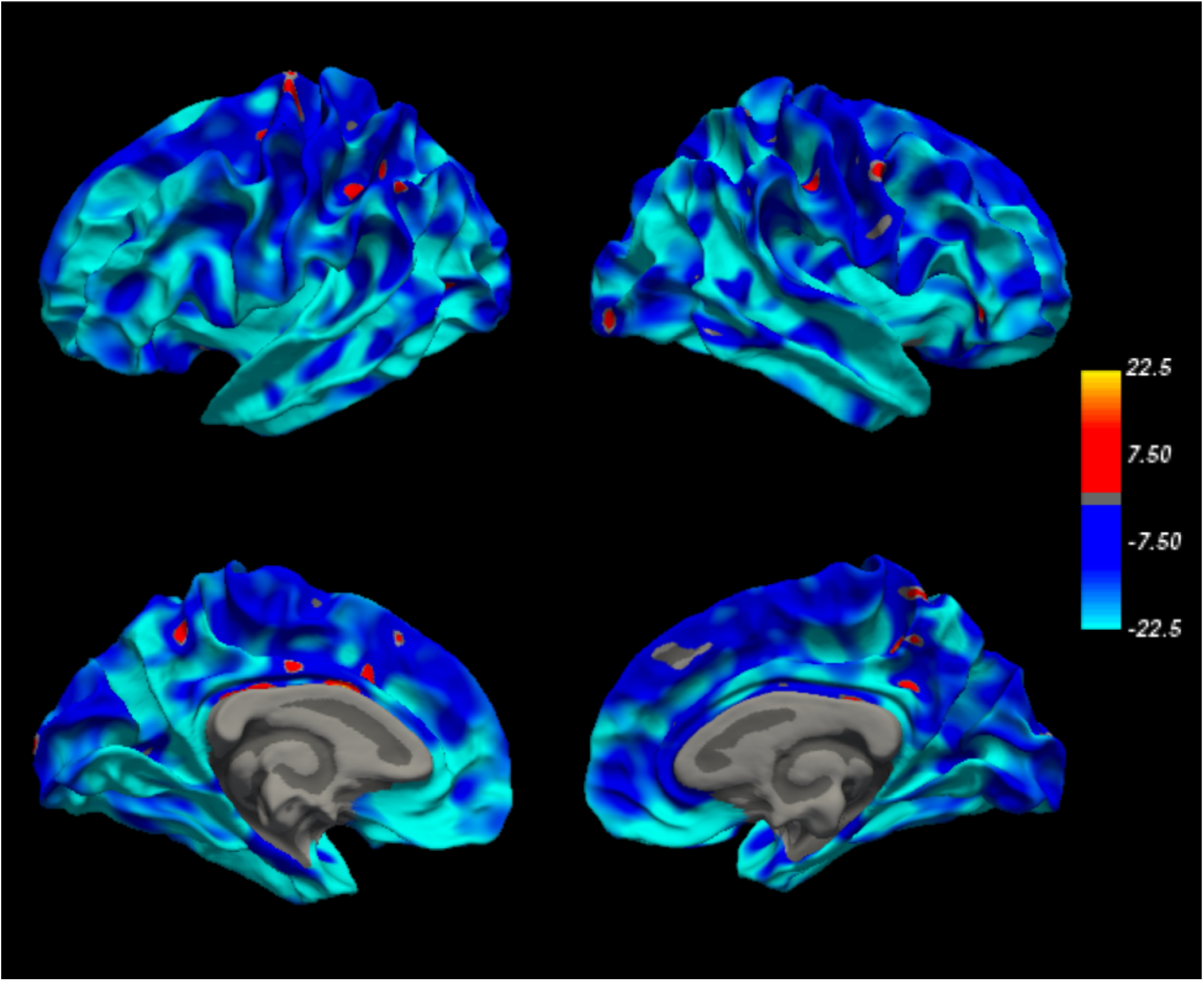
Illustration of the percent difference in neocortical 5-HT2AR binding potential (BPND) between COC users compared to non-users via vertex-based parametric images from the [11C]Cimbi-36-only PET scans. The color bar reflects percent difference between COC users and non-users.

### 5-HT2AR/5-HT2CR binding in subcortical brain regions

Support for the LVM structures was evidenced by significant loadings for each region-specific binding onto the latent variables (all p<0.013). From the score tests, we found no evidence for a lack of fit (P_adj_>0.24), so no additional covariance was added to the models. For the pooled analysis across both radiotracers, we identified no subcortical pattern of an association between COC use and 5-HT2AR/5-HT2CR binding (−7.8%, 95% CI [−21.7;7.7], p=0.31) (**Figure 3A**), however, for the [11C]Cimbi-36-only LVM, we found that COC use was associated with globally lower subcortical 5-HT2AR/5-HT2CR binding (−23.5%, 95% CI [−35.6;−9.1], p=0.002) (**Figure 3B**).

**Figure 3.**
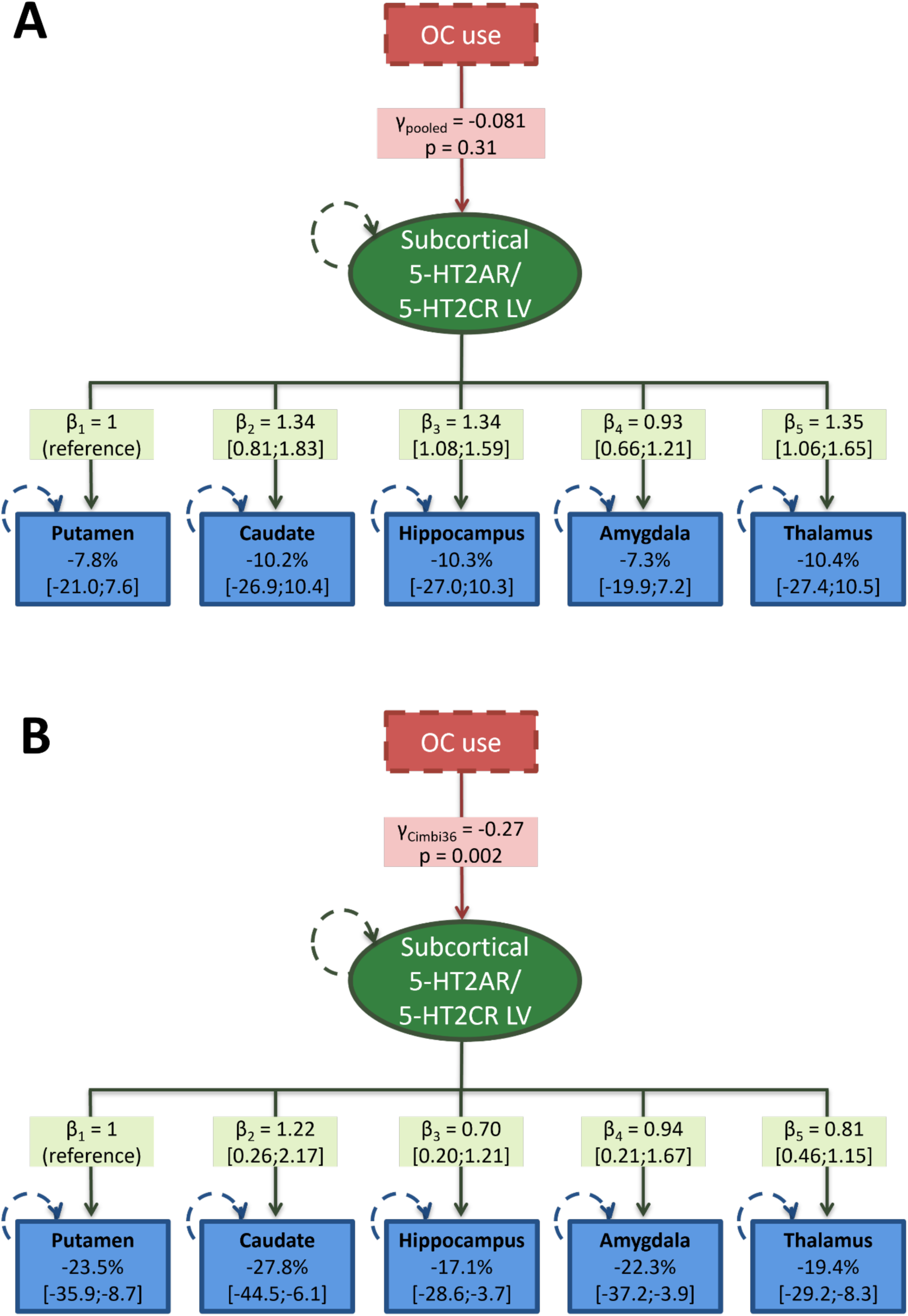
The latent variable models showing the association between combined oral contraceptive (COC) use (represented by the γ-value (on the log-scale)) and the global subcortical 5-HT2AR/5-HT2CR latent variable (LV) (green oval) on the five brain regions (blue boxes) (**A**) in data pooled across [18F]Altanserin and [11C]Cimbi-36 radiotracer scans and (**B**) in [11C]Cimbi-36-only scans. The loading effects on each region are shown as β-values (on the log-scale) with 95% confidence intervals, and the corresponding associated COC effect on the regional 5-HT2AR/5-HT2CR binding potential is shown as percent difference in each blue box. Circular green and blue hatched lines reflect variables estimated with error. Each regional binding potential is independently adjusted for age, body mass index, neuroticism subscore, and for the pooled analysis also radiotracer variant and magnetic resonance field strength (not illustrated). The percent difference in binding potentials reported in the blue boxes was obtained after back transformation using (e^γ·β^ – 1) · 100. Binding potentials could not be estimated for putamen (n=7), caudate (n=3), and amygdala (n=2) due to model fitting issues; imputed values were used as described in the Methods section. **Note:** While [11C]Cimbi-36 binds to both 5-HT2AR and 5-HT2CR subcortically, the [18F]Altanserin binds selectively to 5-HT2AR and has no known affinity for 5-HT2CR.

Differences in the regional loadings led to regional effects ranging from −17.1% (95% CI [−28.6;−3.7]) in the hippocampus, to −27.8% (95% CI [−44.5;−6.1]) in caudate. Sensitivity analyses, including median regression to account for potential outliers are reported in **Table S1**. Again, the results were largely consistent with the main findings, except for twice as low binding in COC users compared to non-users when women with uncertain hormonal contraceptive user status were excluded in the pooled analysis. We found no evidence of a difference in 5-HT2AR binding in subcortical regions between the groups when only including [18F]Altanserin scans (**Figure S3**).

## Discussion

In this cross-sectional study of 71 healthy women, pooling data from both [18F]Altanserin and [11C]Cimbi-36 PET scans, we found no significant group differences in 5-HT2AR and 5-HT2AR/5-HT2CR binding between COC users and non-users across cortical and subcortical brain regions, respectively. However, when restricting our analysis to the 32 women scanned with the [11C]Cimbi-36 radiotracer, we observed lower neocortical 5-HT2AR and lower subcortical 5-HT2AR/5-HT2CR binding in COC users vs non-users, which appeared to be most prominent in the subcortical regions.

As far as we know, only our previous study, in a subgroup of the [18F]Altanserin scanned women presented here, investigated the association between COC use and 5-HT2AR brain levels and found no evidence of an association (Frokjaer et al., 2009). Those findings are now confirmed by our present findings in pooled tracer analyses and the [18F]Altanserin-only subgroup. Thus, the lower binding levels observed with the [11C]Cimbi-36 scans are the first evidence supporting potential COC effects on the 5-HT2AR/5-HT2AR system. Studies on hormone replacement therapy in postmenopausal women have reported increased 5-HT2AR binding following treatment with estrogen, either alone or in combination with progesterone (Kugaya et al., 2003),(Moses-Kolko et al., 2003),(Moses et al., 2000). These findings may appear paradoxical, given that COCs also contain estrogen. However, one possible explanation for the divergent effects between hormone replacement therapy and COCs may be their content of distinctly different types of estrogen. Hormone replacement therapy typically uses natural estrogens such as estradiol, which predominantly activates estrogen receptor beta - highly expressed in serotonergic neurons - whereas COCs usually contain ethinylestradiol, a synthetic estrogen with a stronger affinity for estrogen receptor alpha (Escande et al., 2006). Thus, COC-induced suppression of endogenous estradiol may reduce estrogen receptor beta activation compared to the enhanced signaling seen with hormone replacement therapy (Escande et al., 2006). Alternatively, the progestogen compound as well as differences in hormone dosing, delivery methods, and age-related receptor sensitivity may also contribute to these divergent neurobiological effects of hormone exposures in COC-uses and perimenopausal women.

Beyond 5-HT2AR/5-HT2CR, COCs may exert broader effects on serotonergic signaling. Few studies have been conducted, but one recent cross-sectional study on a closely related serotonin receptor, the 5-HT4R, found that COC use was associated with lower global levels of the 5-HT4R in the brain (Larsen et al., 2020), particularly in regions as the neocortex, amygdala, and hippocampus—areas known to be rich in estrogen and progesterone receptors (Del Río et al., 2018). These results align with the current [11C]Cimbi-36-only findings and are consistent with the notion that COC-induced hormonal changes may modulate postsynaptic serotonergic receptor systems and affect serotonin signaling in ways that may matter for mental health.

From a clinical perspective, the potential reduction in 5-HT2AR/5-HT2CR availability among COC users is noteworthy. The 5-HT2AR is the pharmacological target of both second-generation antipsychotics (5-HT2AR antagonists) and emerging psychedelic-based treatments (5-HT2AR agonists) (Berthoux et al., 2019),(Frokjaer et al., 2008),(Soloff et al., 2010). Moreover, the receptor has also been linked to personality traits and vulnerability to affective disorders (Sankar et al., 2024),(Frokjaer et al., 2008),(Frokjaer et al., 2010). Further, the 5-HT2CR is a potential target in obesity, drug addiction, schizophrenia, and depression [(Higgins et al., 2013),(Lee et al., 2010)] These studies underscore the therapeutic relevance of the 5-HT2AR, highlighting its potential as a target for novel treatments in neuropsychiatric disorders and emphasizing the importance of personalized approaches to enhance treatment efficacy in psychiatric care.

To interpret the diverging results from the [18F]Altanserin and [11C]Cimbi-36 PET radiotracer, it is important to consider both pharmacological and methodological distinctions between the radiotracers. [11C]Cimbi-36 is a 5-HT2AR/5-HT2CR agonist, preferentially binding to receptors in their high-affinity, active state. In contrast, [18F]Altanserin is an antagonist, which binds more broadly—including to inactive receptor pools - potentially reducing specificity (Ettrup et al., 2016). As such, [11C]Cimbi-36 may offer a more accurate reflection of the functional, biologically active receptor pool. Additionally, test-retest variability has been shown to be lower with [11C]Cimbi-36 (<5%) compared to [18F]Altanserin (5–10%) (Ettrup et al., 2016), suggesting greater power to detect changes. Technical differences in PET scanner resolution may also have influenced sensitivity, as [18F]Altanserin scans were acquired using the GE-Advance PET scanner, which has lower spatial resolution than the HRRT scanner used for [11C]Cimbi-36 imaging again adding to noise. Some of Altanserin participants were also scanned using a 1.5T MRI scanner, reducing also the image resolution in the structural scans used for the analysis. Collectively, these factors may have contributed to a reduced signal-to-noise ratio in the [18F]Altanserin dataset, limiting statistical power and increasing susceptibility to Type II error.

Moreover, binding patterns observed with [11C]Cimbi-36 in subcortical regions may reflect combined influences of both the 5-HT2ARs and 5-HT2CRs, rather than 5-HT2AR binding alone. While neocortical regions are largely dominated by 5-HT2AR expression, subcortical areas show a more balanced expression of 5-HT2ARs and 5-HT2CRs (Ettrup et al., 2016),(Ettrup et al., 2014),(Ettrup et al., 2011). Given that [¹¹C]Cimbi-36 binds to both receptors, the observed group differences in subcortical binding may reflect COC-related alterations in either receptor population - or a combination of both. Nevertheless, [¹¹C]Cimbi-36 is expectedly more sensitive to such subcortical group differences than [¹⁸F]Altanserin.

Taken together, these findings highlight the complex and still poorly understood interactions between exogenous sex hormones and central serotonergic systems. Further research is needed to elucidate the receptor- and region-specific mechanisms by which hormonal contraceptives may influence mood and brain function and to understand if hormonal contraception use matter for treatment efficacy of psychiatric disorders.

### Methodological considerations

Although this is, to date, the largest PET study investigating the effects of COC use on 5-HT2AR5-HT2CR levels in healthy women, several limitations must be acknowledged. First, estimating subcortical binding potentials is associated with substantial uncertainty due to the low specific binding of the tracers in these regions, which resulted in modeling failure and therefore missing binding values in some of the regions. Second, the cross-sectional design precludes causal inference. Third, menstrual cycle phase data were unavailable, which prevented adjustment for potential endogenous hormonal fluctuations potentially affecting 5-HT2AR binding (Jamu and Okamoto, 2022),(Pletzer et al., 2018),(De Bondt et al., 2013). Fourth, due to incomplete information on the specific types of COCs used, we were unable to assess whether different formulations and synthetic hormone types may show different association patterns with 5-HT2AR/5-HT2CR binding. Lastly, we assumed that women who reported no current medication use were also not using hormonal contraceptives. However, as hormonal contraception is not always perceived as “medication” in public discourse, this assumption carries a risk of misclassification. Indeed, excluding women with uncertain hormonal contraceptive user status, supported a lower 5-HT2AR/5-HT2CR binding in COC users relative to naturally cycling women.

### Conclusion

This study provides insights into the neurobiological effects of COC use, including potential alterations in 5-HT2AR/5-HT2CR binding. While no significant differences were observed using the antagonist radiotracer, [18F]Altanserin, analyses with the agonist radiotracer, [11C]Cimbi-36. revealed lower neocortical 5-HT2AR binding and lower subcortical 5-HT2AR/5-HT2CR binding in COC users relative to non-users. If replicated, this underscores the importance of considering hormonal contraceptive use and reproductive state in women when evaluating mental health, the efficacy of atypical antipsychotics, and the therapeutic potential of emerging antidepressant treatments targeting the 5-HT2AR/5-HT2CR, including psychedelic compounds.

## Author contributions

Concept and design: A. Kauffmann, V.G. Frokjaer, S.V. Larsen.

Data acquisition/preparation: Kauffmann, A. Sankar, V. Beliveau, V.G. Frokjaer, S.V. Larsen.

Statistical analysis: B. Ozenne, S.V. Larsen, A. Kauffmann.

Interpretation of data: All authors.

Drafting of the manuscript: A. Kauffmann, V.G. Frokjaer, S.V. Larsen.

Critical revision of the manuscript for important intellectual content: All authors.

Obtained funding: V.G. Frokjaer.

Supervision: S.V. Larsen, V.G. Frokjaer.

All authors met the four ICMJE criteria for authorship, read and approved the final manuscript, and agreed to be accountable for all aspects of the work.

## Data Availability

All data produced in the present study are available upon reasonable request to the authors

## Acknowledgement

The authors wish to thank all staff at the Neurobiology Research Unit involved in the collection of data contained within this study, which was conducted using data from the Cimbi database. They also thank the technical staff at the Dept. of Nuclear Medicine, Rigshospitalet for superb technical assistance. Further, they would like to thank the John and Birthe Meyer Foundation for the donation of the Cyclotron and PET scanner as well as the Simon Spies Foundation for the donation of the Siemens Trio MRI scanner.

