## Supplementary material for "Combined oral contraceptive use and serotonin 2A and 2C receptor brain architecture in healthy women"

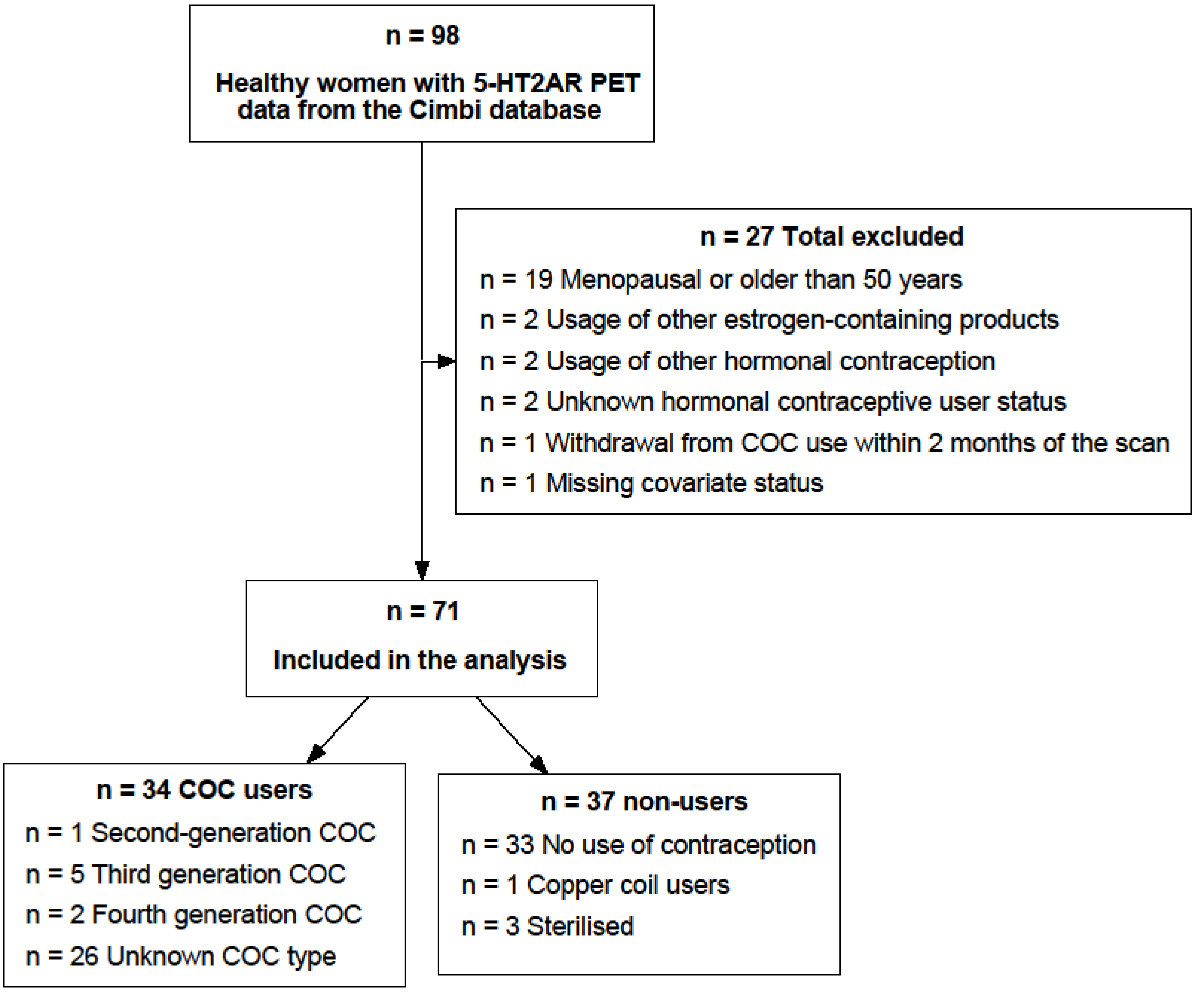


**Figure 1S.** Flowchart of how the study population was derived from the Cimbi database. COC: combined oral contraceptive.

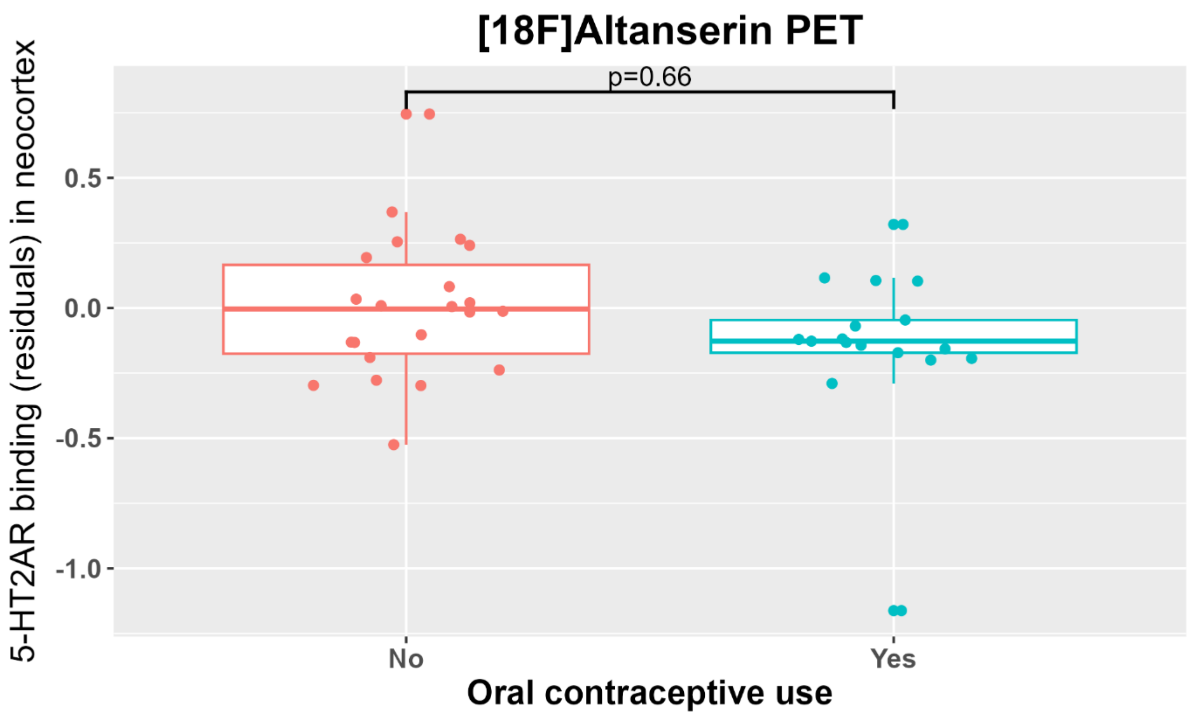


**Figure 2S.** Partial residual plot of log-transformed 5-HT2AR binding potentials in neocortex in COC users and non-users from [18F]Altanserin scans. P-values were computed with multiple linear regression models adjusted for mean-centered age, body mass index, neuroticism subscore, and magnetic resonance field strength (3T as reference).

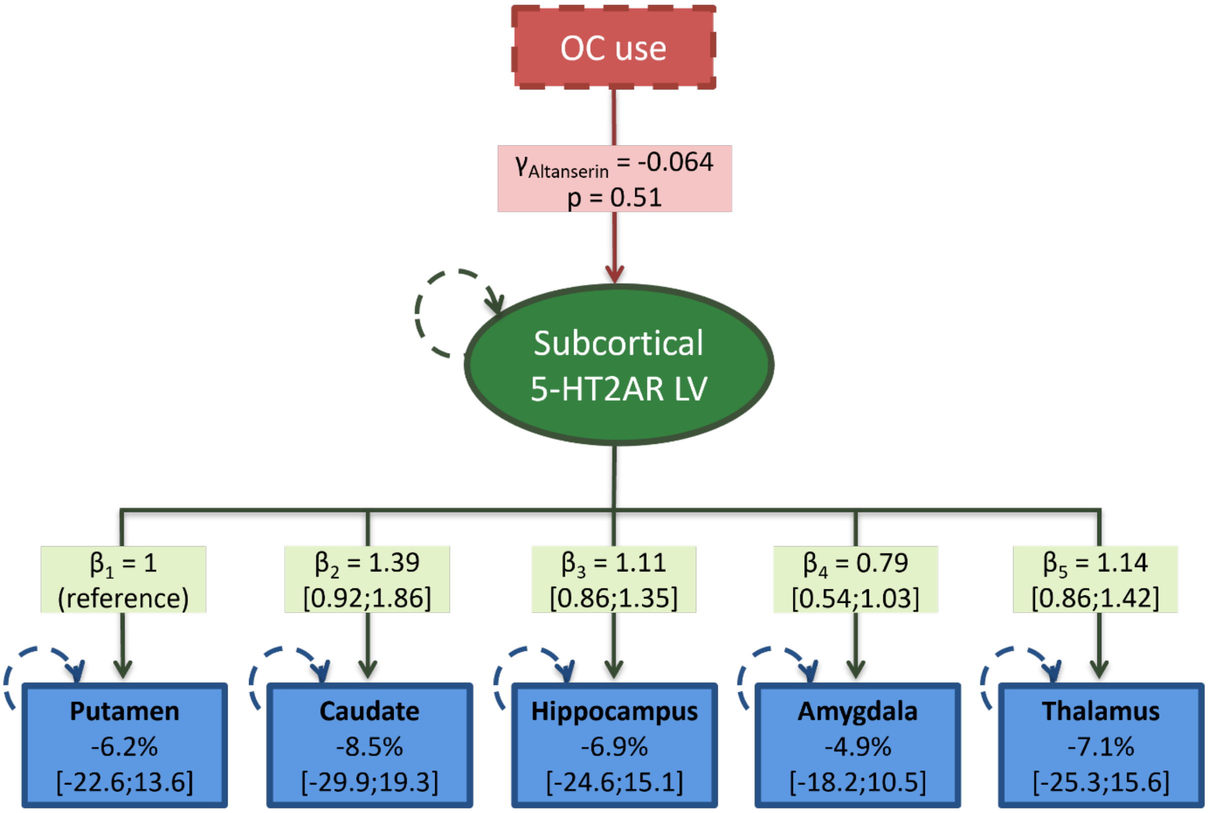


**Figure 3S**. The latent variable models showing the association between combined oral contraceptive (COC) use (represented by the γ-value (on the log-scale)) and the subcortical 5-HT2AR latent variable (LV) (green oval) on the five brain regions (blue boxes) (A) in data from [18F]Altanserin scans. The loading effects on each region are shown as β-values (on the log-scale) with 95% confidence intervals, and the corresponding associated COC effect on the regional 5-HT2AR binding potential is shown as percent difference in each blue box. Circular green and blue hatched lines reflect variables estimated with error. Each regional binding potential is independently adjusted for age, body mass index, neuroticism subscore, and magnetic resonance field strength (not illustrated). The percent difference in binding potentials reported in the blue boxes was obtained after back transformation using (e^(γ·β)-1)·100. Binding potentials were missing for caudate (n=1).

**Table 1S.** Overview of main and sensitivity analyses

| | **Neocortex** | **Subcortical regions** | |
|---|---|---|---|
| | Estimate [95% CI], p-value | Estimate [95% CI], p-value | n |
| **Pooled radiotracers** | | | |
| Main | -7.7% [-18.9; 5.2], p=0.22 | -7.8% [-21.7; 7.7], p=0.31 | 71 |
| Sensitivity 1$^{\alpha}$ | -3.3% [-11.0; 2.9], p=0.63 | -6.3% [-15.4; 3.7], p=0.21 | 71 |
| Sensitivity 2$^{\beta}$ | -12.7% [-24.2; 0.6], p=0.06 | -16.0% [-30.2; 1.2], p=0.07 | 58 |
| **Cimbi-36-only** | | | |
| Main | -12.6% [-22.1; -1.9], p=0.02 | -23.5% [-35.6; -9.1], p=0.002 | 32 |
| Sensitivity 1$^{\alpha}$ | -10.9% [-22.0; 1.4], p=0.17 | - | 32 |
| Sensitivity 2$^{\beta}$ | -16.6% [-25.7; -6.3], p=0.004 | -24.9% [-38.0; -9.1], p=0.003 | 29 |

$^{\alpha}$Median regression. $^{\beta}$Without women with uncertain hormonal contraceptive status
